# Sickle Cell Disease Demographics and Clinical Epidemiology in Gambian Urban and Rural Cohorts Retrospective Analysis

**DOI:** 10.64898/2026.07.03.26357219

**Authors:** Mustapha Dibbasey, Kevin Esoh, Bubacarr Susso, Karen Forrest, Bakary Sonko, Lamin Makalo, Eniyou Oriero, Ndong Ignatius Cheng, Lucas Amenga-Etego, Carla Cerami, Alfred Amambua-Ngwa

## Abstract

Globally, approximately 75% of sickle cell disease (SCD) cases occurs in sub-Saharan Africa, yet empirical data on its natural history, clinical burden, and modifiers remain scarce in the region. This retrospective study describes the demographic characteristics, complications, and routine care and examines how non-genetic factors and blood markers relate to disease severity. We analysed 8402 medical records from 840 SCD patients with confirmed HbSS genotype registered in MRCG Keneba and Fajara clinics (*N*_Keneba_=148; *N*_Fajara_=692). A generalised linear model was employed to estimates association of non-genetic correlates, blood biomarkers, routine care medications with disease severity. Here we showed 67% of patients in Keneba cohort and 92% of those in Fajara cohort had no documented SCD-related chronic complication. Despite no documented evidence of hydroxyurea use, SCD crises (Keneba=0.57, Fajara=0.63) and infections (Keneba=0.53, Fajara=0.35) rates –expressed per patient-year–were low in both cohorts with 99% patients experiencing ≤3 SCD crises per patient-year. Age at diagnosis, gender and seasonality were not significantly associated with SCD crises or other clinical outcomes/events rates. Each additional folic acid prescription was associated with higher haemoglobin(g/dL) (total folic acid prescriptions: β**_Fajara_**=1.31,P=0.005; β_Keneba_=1.20,P<0.001). Penicillin prophylaxis was associated with reduced rate of infection (total Pen V prescriptions: IRR_Fajara_=0.85,P=0.002; IRR_Keneba_=0.93,P=0.002) and SCD crises (IRR_Fajara_=0.67,P=0.001_;_ IRR_Keneba_=0.87,P=0.001). This study found low acute events rates and chronic complications prevalence in the absence hydroxyurea use. No significant associations were observed between non-genetic correlates and clinical events, but the study highlighted the needs for continues folic acid supplementation and penicillin prophylaxis due to their observed beneficial effects.

## Introduction

Sickle cell disease (SCD) is one of the most prevalent monogenic disorders globally, affecting an estimated 300,000 newborns annually, with approximately 75% of cases occurring in sub-Saharan Africa, where early diagnosis and comprehensive management remain suboptimal^1^. SCD comprises a group of disorders, primarily caused by a point mutation in the β-globin gene (*HBB*:c.20A>T) across a genetically heterogeneous background. The single mutation results in the production of sickle haemoglobin (HbS), which polymerizes under low oxygen conditions, leading to erythrocyte deformation, vaso-occlusion, haemolysis, and progressive end-organ damage^2,3^. Two principal pathophysiological processes—vaso-occlusion and haemolysis—underlie the broad spectrum of clinical manifestations observed in individuals with SCD, including acute crises, severe anaemia, chronic and acute organ complications^4–7^.

The clinical phenotype of SCD is notably heterogeneous, ranging from relatively mild symptoms to life-threatening complications such as stroke, chronic kidney disease, acute chest syndrome, and recurrent pain crises^8,9^. In high-income countries, significant improvements in survival and quality of life have been achieved through widespread newborn screening programmes and the implementation of disease-modifying therapies such as hydroxyurea, prophylactic penicillin, and pneumococcal vaccination^9^. However, in low- and middle-income countries (LMICs), including The Gambia, delays in diagnosis, limited access to specialist care, and the scarcity of such disease-modifying therapies pose substantial challenges to effective SCD management. These challenges often contribute to high clinical morbidity and premature mortalities, compounded by a limited understanding of the natural history and clinical epidemiology of SCD^7^.

SCD severity is usually shaped not only by genetic predisposition but also by a complex interplay of non-genetic factors, including sociodemographic status, environmental influences, and health system disparities^10^. These remain poorly characterized in most African populations, considering the continent’s diverse cultural practices, beliefs and social structures as well as ecological differences such as climatic conditions, altitude and nutritional environments^11^. In The Gambia the presence of a robust electronic medical record system at both rural and urban MRCG healthcare facilities, offers a unique opportunity to examine the real-world effects of non-genetic factors such as age at diagnosis, season, and sociodemographic status on the clinical landscape of SCD.

While prior studies in The Gambia have primarily focused on invasive bacterial infection burden and their antimicrobial resistance patterns in SCD patients or evaluated factors associated with quality of life among individuals with SCD^12–14^, there were limited epidemiological characterizations of SCD. Notably, a recent prospective study by Makalo et al. (2025) explored the clinical epidemiology of SCD in Gambian children aged 5–15 years. However, the study was limited in scope, capturing only a year of clinical records and excluded critical age groups, including SCD patients under five and the adult SCD population. Consequently, generalisable empirical evidence regarding the natural clinical progression, disease burden, and modifying factors affecting individuals with SCD in The Gambia across all age groups is undetermined.

To address these gaps, we conducted a retrospective cohort study using detailed medical records from the MRCG Fajara (an urban setting) and Keneba (a rural setting) clinics. In this study, we describe the demographic characteristics of individuals with SCD in these settings, determine clinical phenotypic landscape of SCD using clinical events as proxies of severity, map spectrum of clinical complications, and assess the association of non-genetic correlates and laboratory indices with clinical events. Additionally, we evaluate routine supportive care utilisation such as folic acid, penicillin and anti-malarial prophylaxis in mitigating disease severity. The analysis shows low rate of acute clinical events and prevalence of chronic complications in both cohort in the absence of hydroxyurea, with no associations between age at diagnosis, sex, or season and major clinical events rates. However, more frequent folic acid prescriptions are associated with higher haemoglobin values, and penicillin use is associated with lower rates of infections and fewer SCD crises, suggesting that these simple, low-cost interventions may play an important role in reducing disease burden in this population.

## Methods

### Ethical Considerations

The study received ethical approval from The Gambia Government/MRCG Joint Ethics Committee (Project ID/ethics ref: 31114). Given that all data retrieved from the two databases were anonymized and coded, individual informed consent was waived by the Ethics Committee. Throughout the data access process, participant anonymity was ensured by omitting patient names and exclusively referencing records based only on the West Kiang or MRCG number. All methods were executed in accordance with the Declaration of Helsinki.

### Study Sites, Design and Populations

The MRCG Keneba clinic is a primary healthcare clinic at the MRCG Keneba field station that provides healthcare services to the rural population residing in the Kiang West region. The clinic is equipped to provide routine medical care to patients including SCD patients. MRCG Fajara clinic is within the Kanifing municipality, which is the largest urban setting in coastal western region of The Gambia. MRCG Fajara clinic caters for SCD patients from a variety of backgrounds and regions; however, the majority of SCD patients visiting this clinic originate from the urban centres in The Gambia. The detailed description of both clinics is found in the Supplementary Material file.

This study employed a retrospective design, leveraging the available electronic medical records of SCD patients monitored at both the MRCG Keneba and Fajara clinics. The electronic medical records for SCD patients were captured in the Keneba Electronic Medical Record System (KEMReS) and the MRCG Fajara Clinic Electronic Medical Record System (EMRS) databases. KEMReS has compiled over 15 years of medical records commencing from 2009 and encompassing all age groups^15^. EMRS officially commenced the documentation of medical records for patients attending the MRCG Fajara Clinic in 2016^16^.

### Data Extraction and Variables

Clinical and laboratory data including the genotype data, were retrieved from both KEMReS and EMRS. The data extracted initially relied on the ICD-10 code D57 for sickle cell disorders^17^. The data obtained from both databases were subsequently verified against haemoglobin genotype records in the clinical haematology laboratory department. Consequently, the medical records of only patients with confirmed SCD status (HbSS) were included in subsequent analyses (Figure 1A). The recorded haemoglobin genotyping was determined by alkaline-based haemoglobin electrophoresis method as previously described^5^.

**Figure 1:**
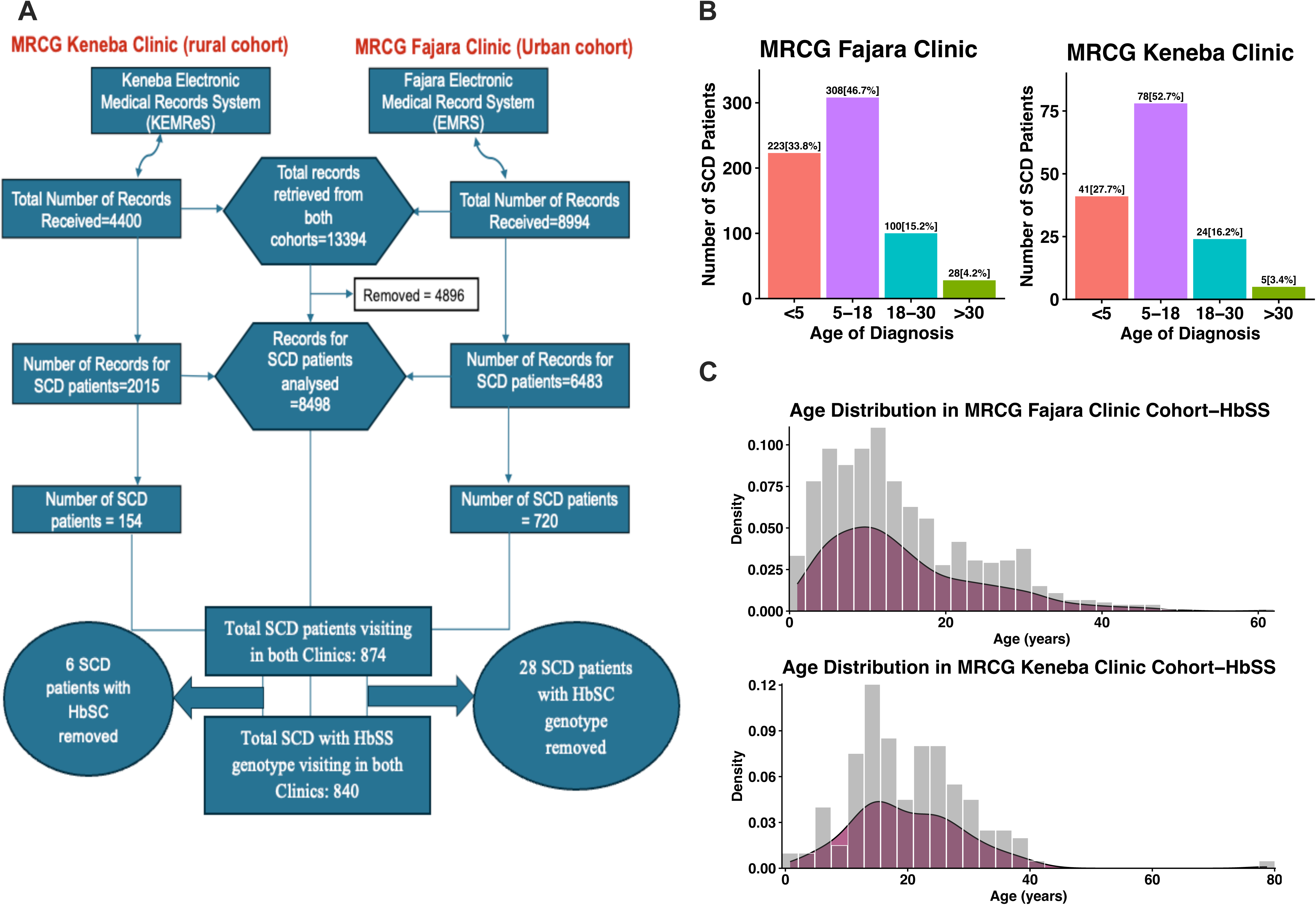
Flow chart, age at diagnosis, age distribution among HbSS patients. A) Flow chart of the data extracted and the number of SCD patients with electronic medical records from the rural and urban cohorts. **B)** The age at diagnosis in the MRCG Keneba Clinic (i.e. in the rural cohort) **and** MRCG Fajara Clinic (i.e. in the urban cohort). C) Age distribution among SCD patients with HbSS genotype

The existing electronic medical data of SCD patients from both the MRCG Keneba and Fajara clinics was descriptively analysed. This included variables on gender, age, age of diagnosis (calculated precisely from date of birth and first laboratory-confirmed SCD diagnosis), ethnicity, past medical history including clinical notes, treatment information, prophylaxis used, and laboratory data on haematological indices such as haemoglobin, total white cell count and differentials and biochemical parameters such as liver function tests, serum creatinine, and urea. Based on the important developmental trajectory of SCD, patients were grouped into different age groups: 0-5; 5-10; 10-18; 18-30; and 30 and above.

### Chronic Complications and Clinical Events

We subsequently examined the documented complications in the clinical data. These complications were identified based on their ICD-10 codes in the electronic medical records supplemented by the clinical notes of the patients that were reviewed electronically. SCD-related complications were categorized into two major classes: acute complications, defined as events necessitating urgent medical intervention or hospitalization (e.g., SCD crises, acute chest syndrome, infections, or acute severe anaemia), and chronic complications, which encompassed long-term comorbidities such as cardiovascular diseases including stroke, avascular necrosis, leg ulcers, and other organ-related damage^9^. Specific clinical outcomes/events known to serve as proxies for disease severity in the clinical course of SCD–Hb-SS disease with crisis with ICD-10 code D57.0 (SCD crises), hospitalizations, infections, and whole blood transfusions ^4,6,18^– were further analysed to determine the clinical severity of SCD **(Table S1).** Acute visits (i.e. clinic visits) were identified by crisis/complication codes regardless of scheduled status due to coding heterogeneity.

### Statistical Analysis

The electronic data from KEMReS and EMRS were retrieved in Excel format and analysed using R version 4.5.1 programming software^19^. The test for normality using Shapiro-Wilk test was validated through Q-Q plots, and a normal distribution bell-shaped curve was employed to visualise and examine the distribution pattern of the data across various clinical parameters and laboratory indices. Continuous variables, such as laboratory data and age at diagnosis, were summarised using descriptive statistics (mean, median, standard deviation, and interquartile range) based on the observed distribution pattern. SCD-related complications were summarized in terms of frequencies (e.g., frequency of SCD/vaso-occlusive crises, hospitalisations, and outpatient clinic visits) and proportions (e.g., the proportion of chronic complications such as stroke in SCD patients). All frequencies of clinical outcomes were adjusted per year, as described previously (Wonkam et al. 2017) and were restricted to SCD patients with more than one year of medical data and at least one clinic visit per calendar year following initial presentation in the databases for outpatient clinic visits.

Age and gender subgroups are critical variables that may modify the natural clinical course of SCD. Consequently, subgroup analyses were conducted based on age groups and gender. Appropriate parametric statistical tests (e.g., t-tests, ANOVA) or their non-parametric counterparts (e.g., Mann-Whitney U test and Kruskal-Wallis test) were employed to compare data across subgroups to identify disparities or trends followed by post-hoc analysis using Dunn test^20^.

The associations between non-genetic correlates (e.g., age at diagnosis, current age, seasonality) and clinical outcomes (e.g., rate of SCD crises, hospitalizations) were assessed using negative binomial regression model where appropriate. Given some variables demonstrated overdispersion, high variable inflation factor, and zero inflation, negative binomial regression model were subsequently employed to derive estimates. The estimates were then converted to incidence rate ratio (IRR) with 95% confidence intervals and p-value^21–23^. The separate forward stepwise negative binomial regression model was used to assess the association between laboratory parameters and all identified clinical outcomes/events, while controlling for age and gender. Similarly, treatment effects on SCD crises were also assessed using negative binomial regression^21,22^. In addition to negative binomial regression, the treatment effects of folic acids on haemoglobin levels were assessed using linear model converted to β estimates with standard errors and 95% confidence interval. Further, we estimated propensity scores–a predicted probability of being in high exposure group (i.e. greater than median propensity score) against low exposure group (i.e. less than or equals to median score)–for each SCD patients as described by^24^. Results were expressed as IRR with 95% confidence intervals and p-values. Only patients with HbSS genotype were included in multivariate modelling of non-genetic correlates. P value of 0.05 was used to determine statistical significance.

## Results

### Demographic chacteristics of the SCD patients in both cohorts

A total of 8498 medical records were retrieved and analysed for 874 SCD patients registered in both MRCG Keneba and Fajara clinic (**Figure 1A**). Only two SCD haemoglobin genotypes statuses, HbSS and HbSC, were encountered in the genotype records, with HbSS representing the largest proportions (96.1%) (**Table 1**). SCD patients with the HbSC genotype were excluded, resulting in a total of 840 SCD patients with the HbSS genotype for further analysis of 8402 medical records **(Figure 1A)**. There were more males than females with SCD across the two cohorts (Keneba cohort [Male: 51.9%], Fajara cohort [Male: 52.2%]).

**Table 1:**
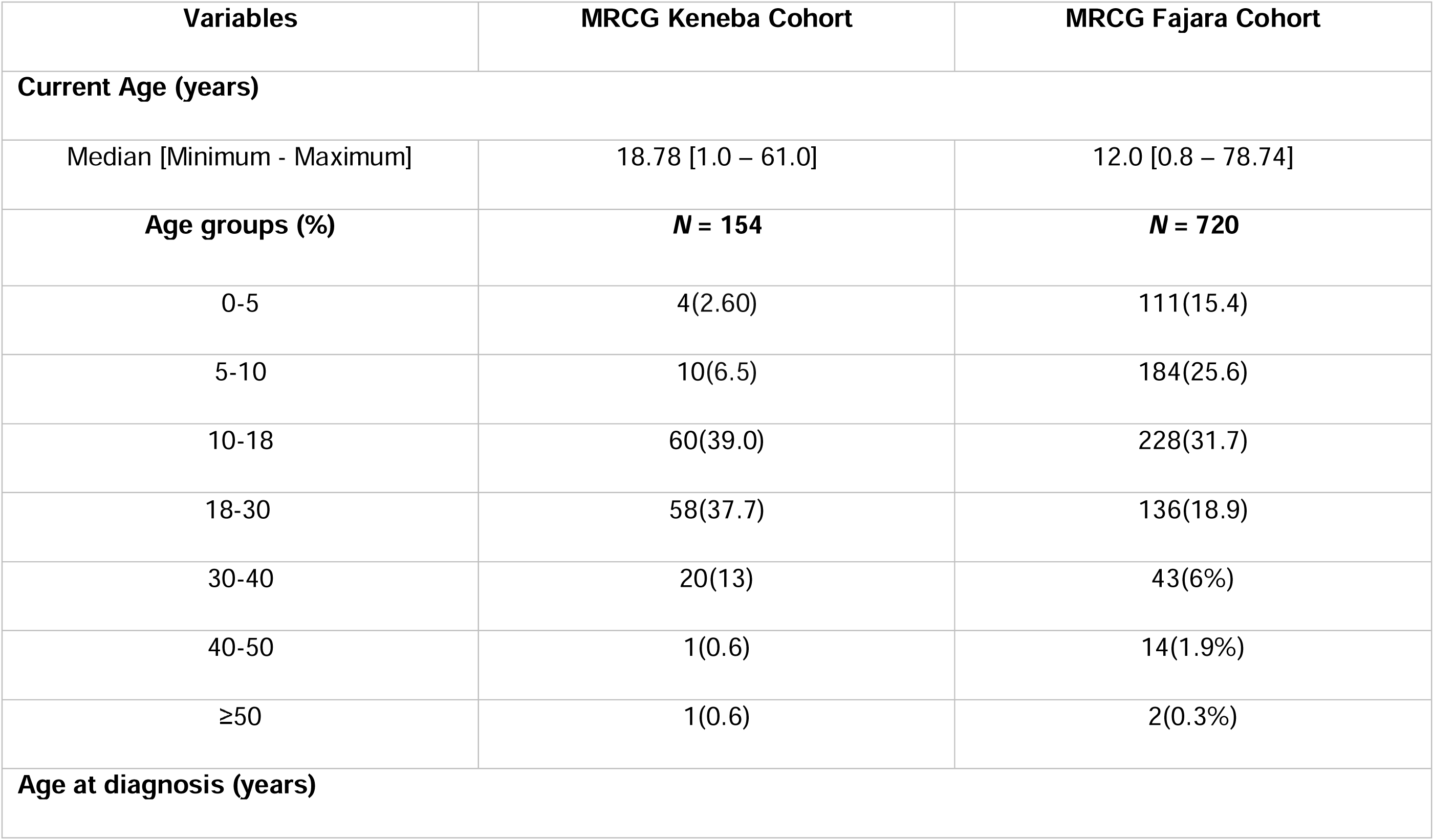
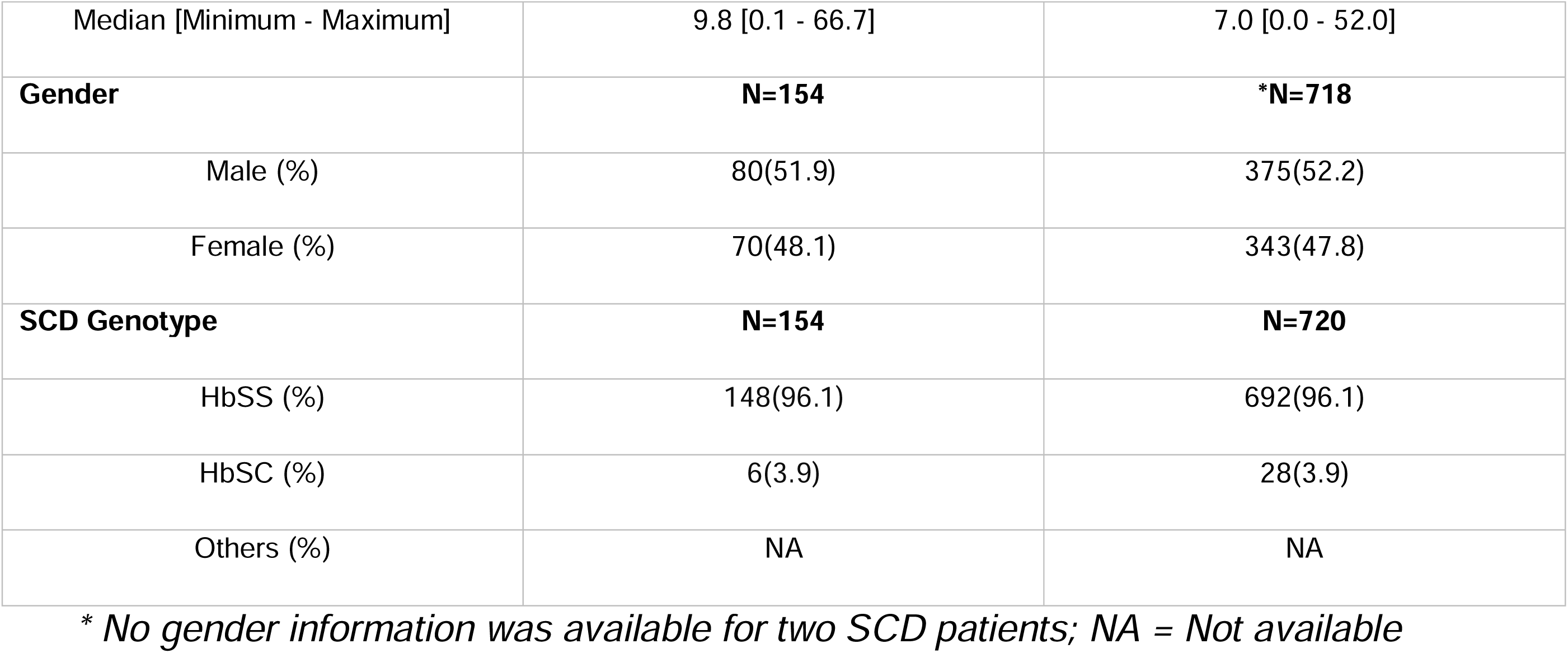
Demography and genotype characteristics of the two cohorts in The Gambia with comprehensive electronic medical records.

Age is a major determinant of clinical manifestation and mortality in SCD^9^. As presented in **Table 1** and **Figure 1C** there was a wide age spectrum (0-80 years) among SCD patients, with median age slightly lower for SCD patients visiting MRCG Fajara clinic (Fajara = 12 years, Keneba = 19 years). We further grouped the patients into age groups based on the developmental trajectory of SCD patients ^25^.

The adolescent age group (10-18) was the most represented in both cohorts (Keneba = 39%, Fajara = 31.7%). Long-term SCD survivors defined as SCD patients surviving beyond 30 years of age in Africa ^26^ were found in considerable proportions (Keneba = 22 [14.2%], Fajara = 59 [8.2%]). Some important laboratory indices showed opposing trends with age in both cohorts (Figure S7 and Figure S8: see the detail in the Supplementary Material file).

Age at diagnosis serves as a crucial risk factor for the severity and morbidity associated with SCD, particularly in LMICs where diagnosis is often established only when a patient presents classical manifestations of the disease such as painful SCD crises^27^. In this study, most patients were diagnosed after 5 years of age (**Figure 1B):** specifically, 9.8 and 7.0 years for Keneba and Fajara respectively **(Table 1).** SCD crises with pain (particularly bone and abdominal pain) and infections were the primary reasons for clinic visits in both cohorts prompting the initiation of SCD diagnosis (**Figure S1)**.

We observed that 7 of the 154 SCD patients from the Keneba clinic were deceased in records retrieved (**Figure S3A and Table S2**). Complications including malnutrition, stroke, leg ulcers, and severe anaemia related to kidney disease were implicated as contributors to mortality among the seven deceased. Records of mortality were not reported in the data retrieved from the Fajara clinic.

### Burden of SCD-related Acute and Chronic Complications

The proportion of individuals experiencing each type of complication was calculated separately for each cohort. SCD crises, infections such as septicaemia, and severe anaemia were identified as the most prevalent acute SCD-related complications in the rural cohort **(Table 2).** The most frequently reported chronic complications of SCD included leg ulcers, stroke and hypertension (**Table 2**).

**Table 2:**
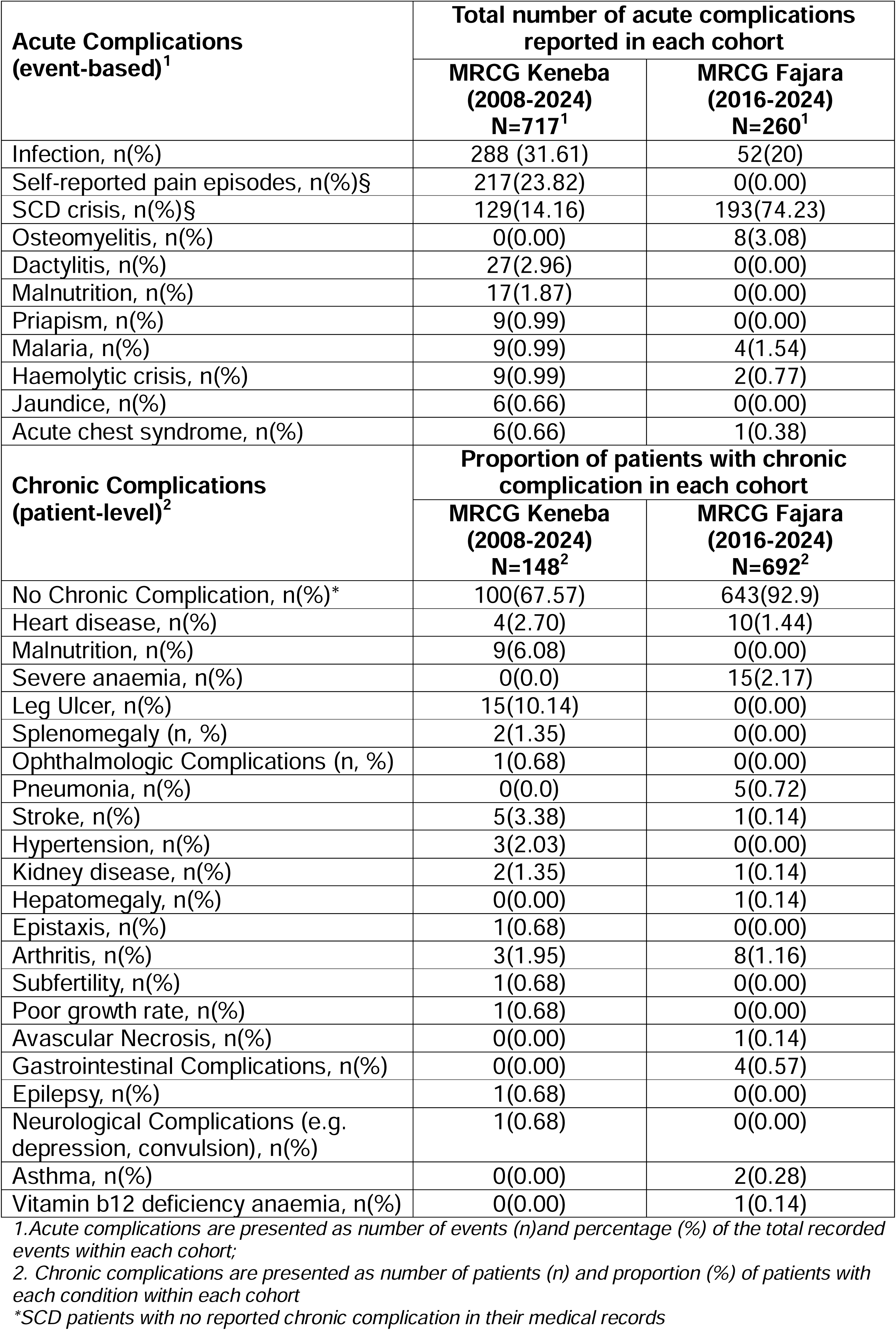
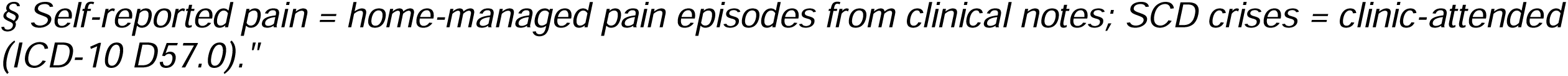
Acute (event-based) and chronic (patient-level) complications in the MRCG Keneba and Fajara SCD cohorts, derived from electronic medical records.

| **Acute Complications**<br>**(event-based)<sup>1</sup>** | **Total number of acute complications**<br>**reported in each cohort** |  |
| --- | --- | --- |
|  | **MRCG Keneba**<br>**(2008-2024)**<br>**N=717<sup>1</sup>** | **MRCG Fajara**<br>**(2016-2024)**<br>**N=260<sup>1</sup>** |
| Infection, n(%) | 288 (31.61) | 52(20) |
| Self-reported pain episodes, n(%)§ | 217(23.82) | 0(0.00) |
| SCD crisis, n(%)§ | 129(14.16) | 193(74.23) |
| Osteomyelitis, n(%) | 0(0.00) | 8(3.08) |
| Dactylitis, n(%) | 27(2.96) | 0(0.00) |
| Malnutrition, n(%) | 17(1.87) | 0(0.00) |
| Priapism, n(%) | 9(0.99) | 0(0.00) |
| Malaria, n(%) | 9(0.99) | 4(1.54) |
| Haemolytic crisis, n(%) | 9(0.99) | 2(0.77) |
| Jaundice, n(%) | 6(0.66) | 0(0.00) |
| Acute chest syndrome, n(%) | 6(0.66) | 1(0.38) |
| **Chronic Complications**<br>**(patient-level)<sup>2</sup>** | **Proportion of patients with chronic**<br>**complication in each cohort** |  |
|  | **MRCG Keneba**<br>**(2008-2024)**<br>**N=148<sup>2</sup>** | **MRCG Fajara**<br>**(2016-2024)**<br>**N=692<sup>2</sup>** |
| No Chronic Complication, n(%)* | 100(67.57) | 643(92.9) |
| Heart disease, n(%) | 4(2.70) | 10(1.44) |
| Malnutrition, n(%) | 9(6.08) | 0(0.00) |
| Severe anaemia, n(%) | 0(0.0) | 15(2.17) |
| Leg Ulcer, n(%) | 15(10.14) | 0(0.00) |
| Splenomegaly (n, %) | 2(1.35) | 0(0.00) |
| Ophthalmologic Complications (n, %) | 1(0.68) | 0(0.00) |
| Pneumonia, n(%) | 0(0.0) | 5(0.72) |
| Stroke, n(%) | 5(3.38) | 1(0.14) |
| Hypertension, n(%) | 3(2.03) | 0(0.00) |
| Kidney disease, n(%) | 2(1.35) | 1(0.14) |
| Hepatomegaly, n(%) | 0(0.00) | 1(0.14) |
| Epistaxis, n(%) | 1(0.68) | 0(0.00) |
| Arthritis, n(%) | 3(1.95) | 8(1.16) |
| Subfertility, n(%) | 1(0.68) | 0(0.00) |
| Poor growth rate, n(%) | 1(0.68) | 0(0.00) |
| Avascular Necrosis, n(%) | 0(0.00) | 1(0.14) |
| Gastrointestinal Complications, n(%) | 0(0.00) | 4(0.57) |
| Epilepsy, n(%) | 1(0.68) | 0(0.00) |
| Neurological Complications (e.g.<br>depression, convulsion), n(%) | 1(0.68) | 0(0.00) |
| Asthma, n(%) | 0(0.00) | 2(0.28) |
| Vitamin b12 deficiency anaemia, n(%) | 0(0.00) | 1(0.14) |

1. Acute complications are presented as number of events (n) and percentage (%) of the total recorded events within each cohort;
2. Chronic complications are presented as number of patients (n) and proportion (%) of patients with each condition within each cohort
\*SCD patients with no reported chronic complication in their medical records
§ Self-reported pain = home-managed pain episodes from clinical notes; SCD crises = clinic-attended (ICD-10 D57.0)."

For the urban cohort, SCD crises, infections such as septicaemia, and severe anaemia were identified as the most prevalent acute SCD-related complications with resultant hospitalisations. For chronic complications, leg ulcer, chronic severe anaemia, heart disease, arthritis, and osteomyelitis were among the most common chronic complications in the SCD patients. Furthermore, there were confirmed incidents of malarial infections in SCD patients: four in the Fajara cohort and nine in the Keneba cohort (**Table 2**). Notably, 92.9% SCD patients (643/692) and 67.6% (100/148) found to have no documented record of chronic complication in the Fajara and Keneba cohort respectively **(Table 2).**

Following descriptive profiling of the study cohorts, we sought to identify clinical events (i.e. outcomes) **(Table S1).** Outcomes were standardized per 100 patient-year to account for variable follow-up durations (i.e. clinic visits), and analysed either as continuous or grouped categorical variables, depending on their distributions.

These clinical events have been utilized as proxies for disease severity in previous SCD research^7,28–30^, especially in the absence of universally validated clinical severity scoring tools. Outcomes were further stratified by cohort (rural and urban) to evaluate regional differences (Table 3) and age groups (Table S3, the detail in the Supplementary Material file). Across both cohorts, the rate of SCD crises and infections were 0.57 and 0.53 for rural cohort and 0.65 and 0.3 for urban cohort, respectively, as shown in **Table 3**. For Fajara cohort, the rate of blood transfusion and hospitalisation were extremely low. Specifically, in the Keneba cohort the median (minimum-maximum) values of SCD crises and infections were 0.26 (0.00–4.0) and 0 (0.00–3.5) per year, while in the Fajara cohort, the values were 0.4 (0.00–6.0) and 0.0 (0.00–11) for SCD crises and infection respectively. The differences in median SCD crises and infection rates were not statistically significant between the two cohorts (**Table 3**).

**Table 3:** Description of clinical outcomes standardised by patient-year.

| Clinical outcomes/events | Mean | Median | Interquartile Range |  | Range (minimum-maximum) |
| --- | --- | --- | --- | --- | --- |
| MRCG Keneba Cohort |  |  |  |  |  |
| Clinical visits | 0.75 | 0.24 | 0.21-0.95 |  | 0.01-6.1 |
| SCD crises | 0.57 | 0.26 | 0.0-1.00 |  | 0.0-4 |
| Infection | 0.53 | 0 | 0.0-1.00 |  | 0.0-3.5 |
| MRCG Fajara Cohort |  |  |  |  |  |
| Clinical visits | 3.14 | 3 | 2.0-4.00 |  | 0-13 |
| SCD crises | 0.63 | 0.4 | 0.0-1.00 |  | 0-6 |
| Infection | 0.35 | 0 | 0 |  | 0-11 |
| Hospitalization* | 0.23 | 0 | 0 |  | 0-4 |
| Transfusion (whole blood)* | 0.08 | 0 | 0 |  | 0-2 |
| Comparing the two cohorts (MRCG Keneba vs MRCG Fajara Cohort) | MRCG Keneba Cohort |  | MRCG Fajara Cohort |  | Statistical Analysis (P value)<sup>§</sup> |
|  | Mean | Median | Mean | Median |  |
| Infection | 0.53 | 0 | 0.35 | 0 | - |
| SCD crises | 0.57 | 0.26 | 0.63 | 0.4 | P>0.05 |

\* Clinical outcomes not found in the KEMReS database, Keneba cohort database; <sup>§</sup> Mann Whitney U test (two-sided) statistical analysis performed on the median values

To assess disease severity distributions, SCD patients were grouped based on the number of documented SCD crises per year (0, 1, 2, 3, or ≥3 episodes). As shown in **Figure 2A-D**, most patients experienced either zero (Keneba cohort: *N* = 83; Fajara cohort: *N* = 363) or only one crisis (Keneba cohort: *N* = 73; Fajara cohort: *N* = 259) per patient-year. Notably, only 2 patients in the Fajara cohort and 1 patient in the Keneba cohort experienced more than three SCD crisis episodes per patient-year, representing less than 1% of individuals in each cohort. Hence, more than 95% of SCD patients experienced less than three episodes of SCD crises in both cohorts (**Figure 2A and 2D**). This finding suggests clinical phenotypes among patients in the two cohorts in The Gambia are predominantly mild. Importantly, the observed clinical patterns may reflect the natural history of SCD in the two cohorts, particularly given the complete absence of hydroxyurea therapy across both cohorts (**Figure 3**). Similar findings were also documented in other clinical outcomes such as rate of infection, hospitalisation and transfusion (**Figure 2B and 2D, Figure S6A and S6B**). Information on alternative sources of hydroxyurea treatment for these patients were not available in the records.

**Figure 2:**
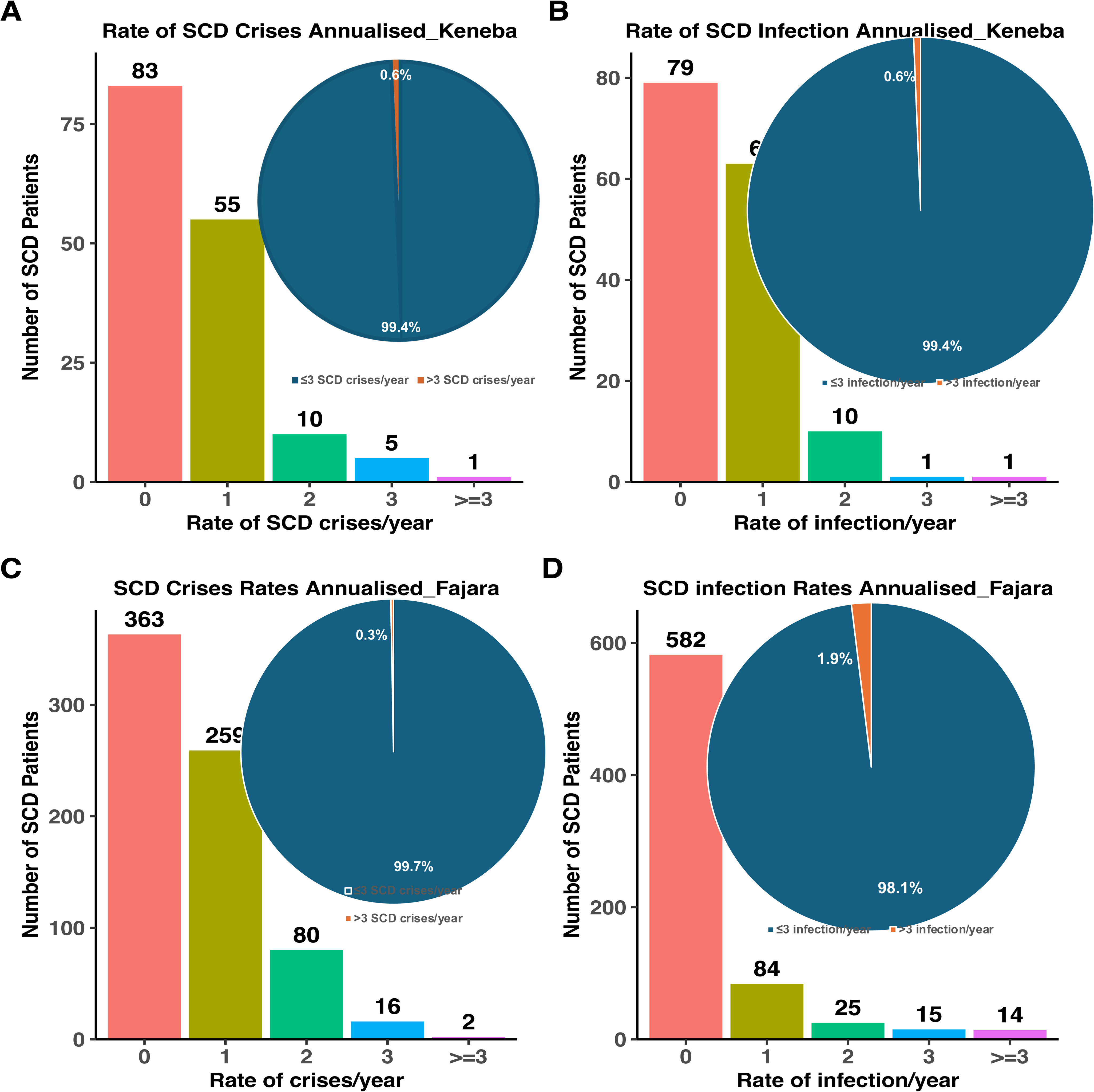
Clinical outcomes of SCD in Keneba and Fajara. **A)** Rate of SCD crises adjusted by patient-year and the proportion of patients with ≤3 and >3 SCD crises/year in rural cohort. **B)** Rate of infection per year adjusted by patient-year and the proportion of patients with ≤3 and >3 SCD crises/year in rural cohort. **C)** Rate of SCD crises adjusted by per patient-year and the proportion of patients with ≤3 and >3 SCD crises/year in urban cohort. **D)** Rate of infection/year adjusted by per patient-year and the proportion of patients with ≤3 and >3 SCD crises/year in urban cohort

**Figure 3:**
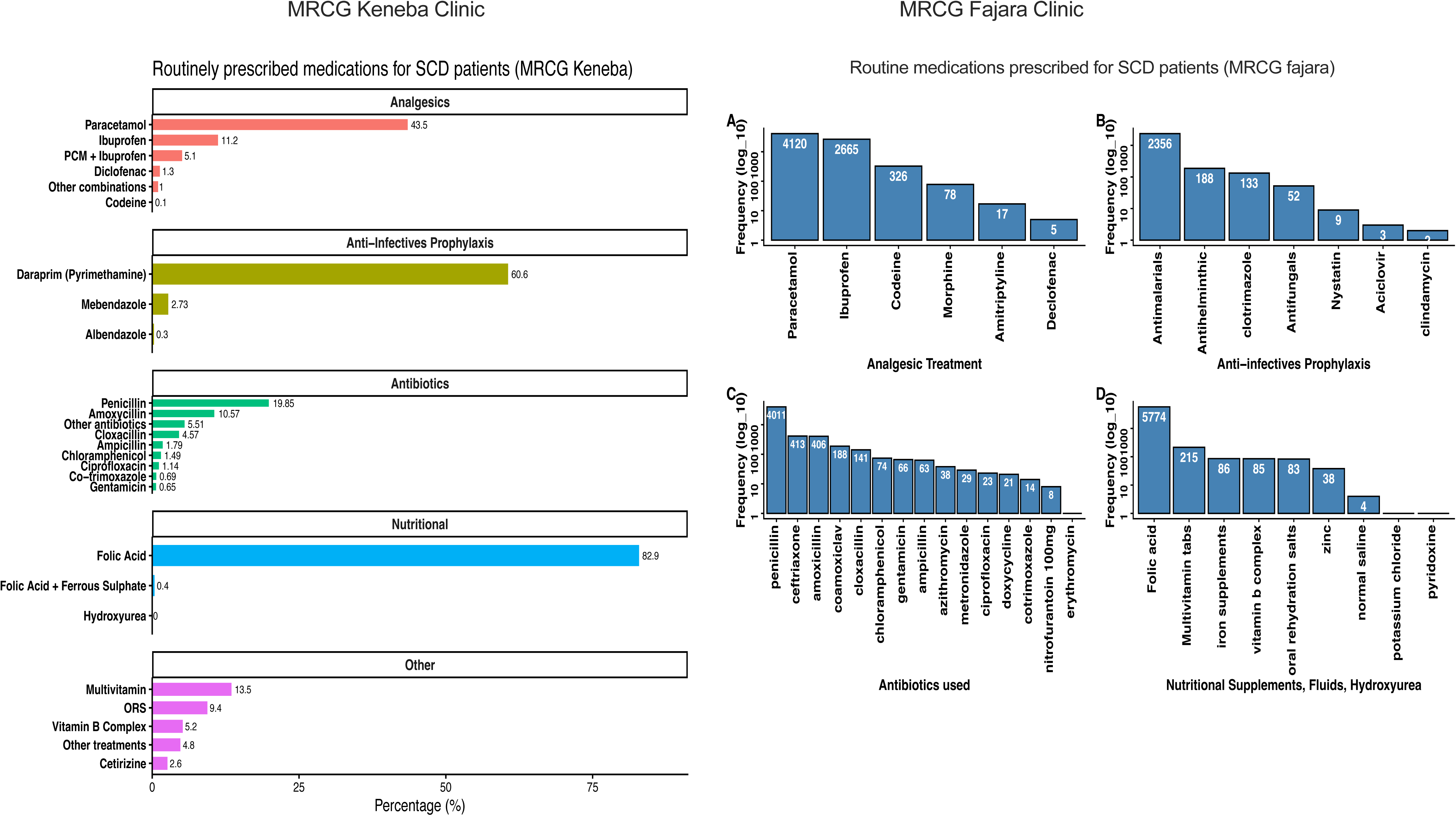
Drugs routinely prescribed for supportive and preventive management of SCD patients in both cohorts. **Left panel (MRCG Keneba Clinic):** Drugs routinely prescribed for supportive and preventive management of SCD patients. **Right panel (MRCG Fajara Clinic): A)** analgesic treatments for pain management **B)** anti-infective prophylaxis for prevention of malaria and fungal **C)** antibiotics used as prophylaxis and for treatment of infections **D)** Nutritional supplements, fluids and hydroxyurea. Hydroxyurea frequency was zero, hence not reported. The numbers in the bar represent the frequency of prescription of each drug in the medical records received.

### Supportive Care Utilisation and Clinical Impacts

Consistent with current standards of comprehensive care in LMICs, all SCD patients routinely received supportive management, including folic acid supplementation for anaemia, and paracetamol and other analgesics for pain management **(Figure 3).** Additionally, antibiotics such as penicillin and amoxicillin were administered either prophylactically for infection prevention in individuals with SCD or therapeutically for active infections in both cohorts (**Figure 3**). Based on the data from both cohorts, we observed that penicillin use was not restricted to only under-five SCD patients (**Figure S4 and Figure S5**). Moreover, there were records of anti-malarial drugs use such as daraprim and mefloquine for prophylactic purpose in both cohorts. Anti-helminthic prophylactics (mebendazole and albendazole) were used in both cohorts. There was no recorded hydroxyurea treatment in both cohorts for the period captured in this study (**Figure 3**).

Having adjusted for potential confounders such as age and sex as well as baseline haemoglobin level, our multivariable regression models indicated that folic acid supplementation was significantly associated with improved haemoglobin levels (g/dL) in both cohorts (**Figure 4)**. Penicillin use was associated with a reduction in infection rates in both cohorts but did not demonstrate a direct effect on the rate of SCD crises in Fajara cohort even though infections were found to be a major precipitating factor for SCD crises leading to hospitalisations **(Table S4).** To further investigate this association, we applied a propensity score analysis in which SCD patients in penicillin exposure group (e.g., those receiving penicillin) were matched with similar SCD patients in the other non-penicillin exposure group (e.g., those not receiving penicillin) by their propensity scores^24^. This additional analysis demonstrated that penicillin use exerted an indirect effect in reducing the rate of SCD crises (IRR: 0.67, 95% CI: 0.50–0.90, P < 0.001) **(Figure 4).**

**Figure 4:**
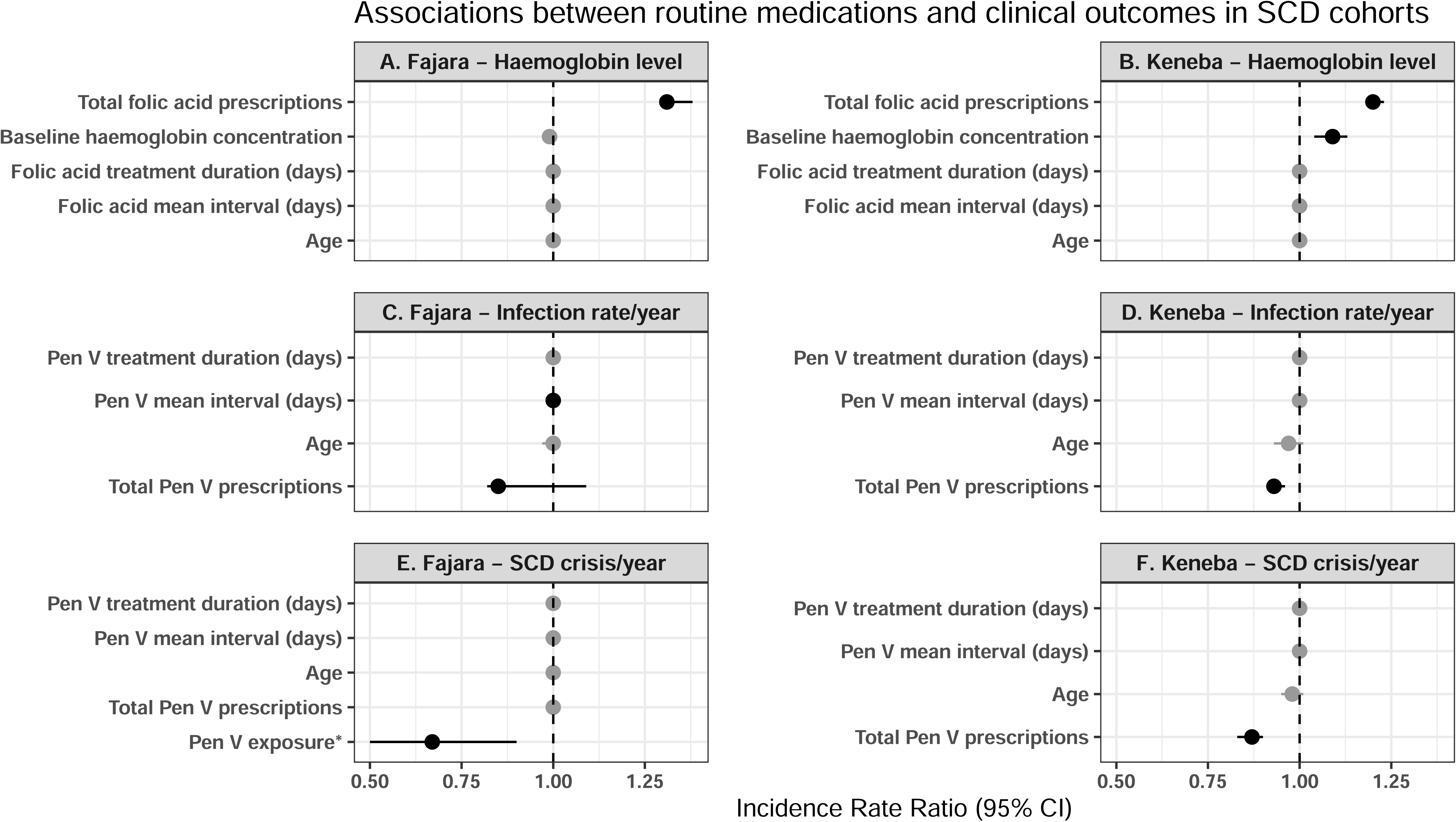
Effects of folic acid and penicillin V prescriptions on SCD outcomes in Fajara (n=692) and Keneba (n=148) cohorts. **(A-B)** Association between number of folic acid prescriptions and haemoglobin (g/dL) in Fajara (A) and Keneba (B) cohorts, analysed using linear regression adjusted for age and baseline haemoglobin concentration (g/dL). The results shown as β estimates with 95% confidence interval (error bars). (C-F) Association between penicillin V prescriptions and infection rate (C-D) or SCD crisis rate (E-F) in Fajara and Keneba cohorts, analysed using negative binomial regression adjusted for age. Results shown as adjusted incidence rate ratios (IRR) with 95% confidence intervals (error bars). \**Pen V exposure in Fajara cohort: propensity score matching performed to group SCD patients based on penicillin exposure and non-exposure; results expressed as* IRR with 95% confidence intervals (error bars).

### Potential Non-genetic Correlates of SCD Severity

To explore the association of non-genetic correlates– age at diagnosis, age, gender, visit month (seasonality), and geography–with the clinical outcomes in the two cohorts, we conducted a multivariable regression analysis. Despite observed heterogeneity in the frequencies of clinical outcomes across age groups and seasonal months, our analysis revealed that age, age at diagnosis, gender, and seasonality did not significantly influence the rate of SCD crisis in either the rural or urban cohorts **(Table 4 and Table 5, Figure S2)**. However, distance from the MRCG Keneba clinic—a key healthcare access point in the rural cohort—was inversely associated with SCD crises rate. Specifically, individuals residing farther from the clinic observed to have a significantly lower risk of SCD crises (Incidence Rate Ratio [IRR]: 0.95; 95% [CI]: 0.90–0.99; P value = 0.024). Non-KWR residents demonstrated nearly threefold higher rates of clinic visits compared to SCD patients residing in KWR (IRR: 2.89; [CI]: *P* = 0.001). In contrast, none of the assessed non-genetic correlates significantly predicted infection rates in the Keneba cohort.

**Table 4:** Non-genetic correlates and laboratory biomarkers association with clinical event in the rural cohort–Keneba clinic based on negative binomial regression model.

| Non-Genetic Correlates of SCD | Clinical outcomes (Estimates (P value)) |  |  |  |  |  |  |  |  |
| --- | --- | --- | --- | --- | --- | --- | --- | --- | --- |
|  | Clinic Visit Rate/year |  |  | SCD Crisis Rate/year |  |  | Infection Rate/year |  |  |
|  | Adjusted IRRs | 95% CI | P value | Adjusted IRR | 95% CI | P value | Adjusted IRR | 95% CI | P value |
| Age at Diagnosis | 0.67 | 0.50-0.85 | **0.01** | 0.99 | 0.88-1.10 | 0.787 | 0.99 | 0.73-16.69 | 0.110 |
| Age (current age) | 1.34 | 1.12-1.64 | **0.01** | 0.99 | 0.89-1.11 | 0.879 | 1.01 | 0.92-1.11 | 0.821 |
| Distance from the Clinic | 0.98 | 0.94-1.01 | 0.10 | 0.95 | 0.90-0.99 | **0.024** | 0.99 | 0.95-1.03 | 0.564 |
| Gender(Female) | Ref |  |  | ref |  |  | ref |  |  |
| Male | 0.71 | 0.48-1.06 | 0.09 | 1.119 | 0.53-2.66 | 0.238 | 0.68 | 0.37-1.25 | 0.223 |
| Ethnicity(Fula) | Ref |  |  | ref |  |  | ref |  |  |
| Mandinka | 1.79 | 0.22-9.42 | 0.52 | 0.56 | 0.11-2.03 | 0.431 | 0.91 | 0.24-2.68 | 0.879 |
| Jola | 0.93 | 0.01-12.27 | 0.48 | 1.33 | 0.14-18.97 | 0.809 | 1.46 | 0.20-13.89 | 0.712 |
| Resident in KWR | ref |  |  | ref |  |  | ref |  |  |
| Non-Resident | 2.89 | 1.82-4.71 | **0.001** | 1.28 | 0.68-3.74 | 0.283 | 1.55 | 0.82-2.97 | 0.183 |
| Season (Dry) | ref | - | - | ref |  |  | ref |  |  |
| Wet | 0.00 | 0.00-0.99 | 0.99 | 1.05 | 0.80-1.38 | 0.702 | 1.05 | 0.79-1.40 | 0.713 |
| **Laboratory Parameters** |  |  |  |  |  |  |  |  |  |
| White Blood Cells | 1.03 | 0.99-1.09 | **0.01** | 1.04 | 0.99-1.11 | **0.032** | 1.04 | 0.99-1.10 | **0.025** |
| Haemoglobin | 1.23 | 0.82-1.86 | 0.28 | 2.56 | 0.91-9.46 | 0.195 | 0.97 | 0.69-1.36 | 0.824 |
| Platelets | 1.00 | 1.0-1.0 | 0.15 | 1.00 | 0.99-1.00 | 0.073 | 1.00 | 1.00-1.00 | 0.371 |
| Red blood cells | 0.46 | 0.18-1.06 | 0.08 | 0.09 | 0.00-1.49 | 0.195 | 0.14 | 0.01-1.00 | 0.149 |
| Mean Cell Volume | 0.96 | 0.92-1.01 | 1.101 | 0.91 | 0.78-1.03 | 0.209 | 0.94 | 0.83-1.02 | 0.258 |
| Mean Cell Haemoglobin Concentration | 0.97 | 0.85-1.11 | 0.69 | 0.84 | 0.62-1.11 | 0.303 | 0.87 | 0.68-1.08 | 0.273 |
| Neutrophils | 1.01 | 0.99-1.03 | 0.13 | 1.04 | 1.01-1.08 | **0.002** | 1.01 | 0.99-1.03 | 0.365 |
| Lymphocytes | 0.99 | 0.97-1.02 | 0.46 | 0.98 | 0.94-1.02 | 0.412 | 0.99 | 0.96-1.02 | 0.548 |
| Monocytes | 0.86 | 0.73-1.01 | 0.06 | 0.82 | 0.59-1.10 | 0.174 | 0.85 | 0.67-1.07 | 0.120 |

*CI: confidence interval; IRR: Incidence Rate Ratio. Statistical significance ( $p < 0.05$ ) and IRR* *values shown in bold. All IRRs adjusted for covariates. No multiple comparison correction* *applied.*

**Table 5:** Non-genetic correlates and laboratory biomarkers association with clinical events in the urban cohort–Fajara clinic based on negative binomial regression model.

| Predictors | Clinic Visit Rate |  |  | SCD Crises Rate/year |  |  | Transfusion Rate/year |  |  | Admission Rate/year |  |  |
| --- | --- | --- | --- | --- | --- | --- | --- | --- | --- | --- | --- | --- |
|  | Adjusted IRR | 95% CI | P value | Adjusted IRR | 95% CI | P value | Adjusted IRR | 95% CI | P value | Adjusted IRR | 95% CI | P value |
| age at diagnosis | **0.98** | 0.96 – 1.00 | **0.027** | 1.00 | 0.99 – 1.01 | 0.437 | **1.14** | 1.01 – 1.31 | **0.039** | 1.03 | 0.97 – 1.11 | 0.327 |
| Current age | 1.01 | 0.99 – 1.02 | 0.396 | **0.99** | 0.98 – 1.01 | **0.044** | **0.85** | 0.74 – 0.95 | **0.007** | 0.98 | 0.91 – 1.04 | 0.465 |
| sex [M] | 0.95 | 0.87 – 1.03 | 0.196 | 1.76 | 0.99 – 3.54 | 0.077 | 1.83 | 1.03 – 3.36 | **0.045** | 1.09 | 0.79 – 1.51 | 0.598 |
| season [Wet] | **1.09** | 1.01 – 1.19 | **0.037** | 1.07 | 0.89 – 1.29 | 0.452 | 1.56 | 0.33 – 27.94 | 0.662 | 0.86 | 0.63 – 1.18 | 0.353 |
| Source [Keneba] | **0.24** | 0.20 – 0.29 | **<0.001** | 0.92 | 0.72 – 1.14 | 0.445 | - | - | - | - | - | - |
| Laboratory Biomarkers |  |  |  |  |  |  |  |  |  |  |  |  |
| Haemoglobin | 1.02 | 0.98 – 1.06 | 0.300 | 0.99 | 0.92 – 1.07 | 0.825 | 0.84 | 0.68 – 1.03 | 0.099 | 0.93 | 0.82 – 1.06 | 0.312 |
| Total white cell count | 1.00 | 0.98 – 1.03 | 0.827 | 0.98 | 0.94 – 1.04 | 0.536 | 0.99 | 0.88 – 1.15 | 0.910 | 1.01 | 0.93 – 1.12 | 0.836 |
| Mean cell volume | 1.00 | 0.99 – 1.00 | 0.783 | 1.00 | 0.99 – 1.01 | 0.873 | 0.97 | 0.94 – 1.00 | 0.095 | 1.00 | 0.99 – 1.02 | 0.614 |
| Mean cell haemoglobin concentration | 1.00 | 0.98 – 1.01 | 0.632 | 1.00 | 0.96 – 1.02 | 0.743 | 1.01 | 0.94 – 1.05 | 0.596 | 0.95 | 0.89 – 1.01 | 0.153 |
| Neutrophils (%) | 1.00 | 0.99 – 1.01 | 0.855 | 1.01 | 0.99 – 1.03 | 0.535 | 0.96 | 0.92 – 1.02 | 0.154 | 0.98 | 0.95 – 1.02 | 0.314 |
| neutrophil absolute | 1.00 | 0.97 – 1.02 | 0.855 | 1.02 | 0.96 – 1.06 | 0.418 | 1.00 | 0.85 – 1.09 | 0.941 | 0.99 | 0.89 – 1.07 | 0.876 |
| Lymphocytes (%) | 1.00 | 0.99 – 1.01 | 0.912 | 0.99 | 0.96 – 1.02 | 0.413 | 0.93 | 0.86 – 1.00 | **0.035** | 0.94 | 0.90 – 0.99 | **0.022** |
| lymphocyte absolute | 1.02 | 0.97 – 1.08 | 0.352 | 1.08 | 0.98 – 1.19 | 0.104 | 1.16 | 0.89 – 1.48 | 0.258 | 1.09 | 0.92 – 1.28 | 0.308 |

CI: confidence interval; IRR: Incidence Rate Ratio. Statistical significance ( $p < 0.05$ ) and IRR values shown in bold. All IRRs adjusted for covariates. No multiple comparison correction applied.

In the Fajara cohort, we observed modest associations of SCD crises (IRR: 0.99; 95% CI: 0.98–1.00; *P* = 0.044) and blood transfusion (IRR: 0.85; 95% CI: 0.74–0.95; *P* = 0.007) with age, indicating patients with fewer crises and blood transfusion rates tended to be older than patients who had more crises and higher rates of transfusion **(Table 5).** Besides, age at diagnosis was associated with an increased blood transfusion rate for severe anaemia treatment (IRR: 1.14; 95% CI: 1.01–1.31; *P* = 0.039).

We also examined the seasonal distribution pattern of SCD crises relative to climatological patterns in rural-KWR and urban centres-Serekunda (**Figure S6).** In the Fajara cohort, although seasonality was not statistically significantly associated with rate of SCD crises as shown in **Table 5**, the temporal trend in SCD crises events **(Figure 5B)** mirrored the country’s humidity profile of Serekunda region as shown in **Figure S6A**. In the Keneba cohort, SCD crises peaked in April and declined to their lowest in October **(Figure 5A)**, albeit with no clear correlation with temperature and humidity pattern from KWR shown in **Figure S6B.**

**Figure 5:**
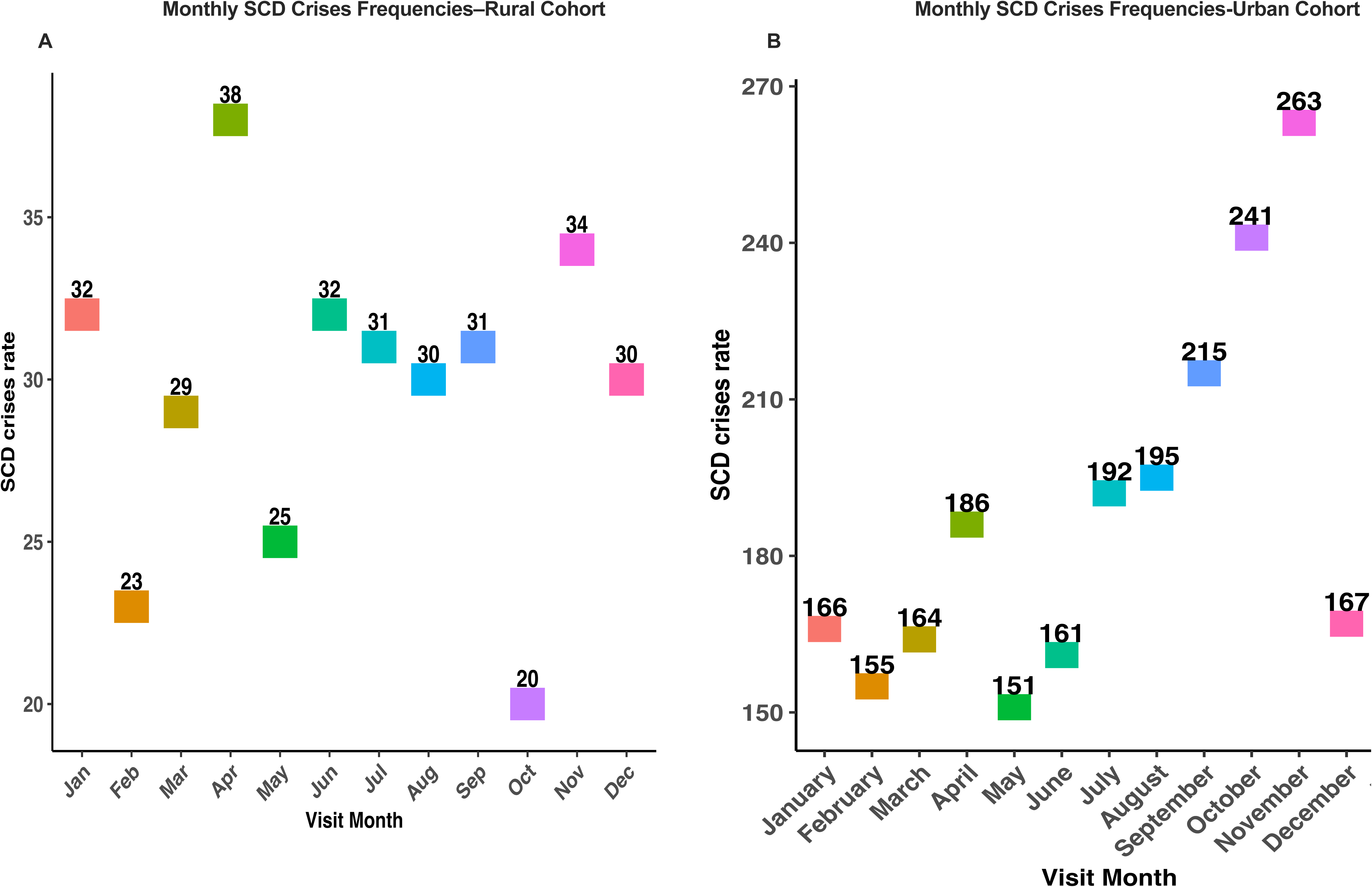
Seasonal effects: Monthly reported frequency of SCD crises in both cohorts. (**A)** Rural cohort: sickle cell disease crises data extracted from MRCG Keneba clinic database, KEMReS. (**B)** Urban cohort: sickle cell disease crises data extracted from MRCG Fajara clinic electronic medical records system database.

## Discussion

This study represents one of the first large-scale analyses of clinical data of SCD patients in The Gambia and contributes essential phenotypic data to support improved clinical management. It also establishes a foundation for future research into genetic modifiers of SCD severity in The Gambia and provide data on SCD clinical epidemiology that fit within the broader framework of data sharing with other African countries and the SickleInAfrica consortium with the aim to inform African-specific future research directions on SCD^31^. Our findings offer critical insights into the demographic characteristics and clinical phenotype landscape, burden of SCD-related clinical complications, effectiveness of existing supportive management strategies for SCD patients, and non-genetic correlates of SCD severity in a sub-Saharan African country where such data remain scarce.

The first five years of life represents a critical period in the clinical course of patients with SCD, during which 90–95% of affected children in sub-Saharan Africa die prematurely^32^. The demographic characteristics of SCD population revealed two interesting findings: a small but significant proportion of SCD patients surviving beyond 30s who are otherwise called long-term survivors and the presence of delayed diagnosis, which is common in countries such as the Gambia where universal newborn screening programmes are yet to be implemented^9^. The existence of long-term survivors provides a valuable opportunity to investigate the socio-demographic and genetic factors that underpin prolonged survival in resource-limited settings. In contrast, delayed diagnosis deprives affected individuals of timely interventions, including penicillin and antimalarial prophylaxis, and hydroxyurea therapy. The most common triggers for SCD diagnosis were pain crises and infections, which often reflect advanced disease presentations and consistent with findings from cooperative study of SCD (CSSD) cohort ^28^. This delayed diagnosis likely contributed to the underestimation of SCD burden in early life in both cohorts and could limit the potential for early intervention, as emphasized in other African studies^33,34^. These findings underscore the critical importance of implementing universal newborn screening programmes for SCD in the Gambia that would lead to early and timely SCD diagnosis. Early diagnosis enables the timely initiation of prophylactic and therapeutic interventions, facilitating comprehensive care and significantly improving clinical outcomes while reducing morbidity and mortality.

The majority of SCD patients experienced less than three SCD crises per year, alongside low rate of hospitalization, transfusion, and infection which suggest milder SCD phenotypes predominates in The Gambia and is aligned with a recent publication from The Gambia that focused on only SCD patients from 5 to 15 years of age^35^. Our finding is also consistent with report from Senegal and India^5,18,26,29,36^. For instance, a study by Seck et al. 2024 in a Senegalese cohort found that over 95% of SCD patients experienced fewer than three SCD crises per year and more than 50% of SCD patients had no documented SCD crises when adjusted per patient-year. In contrast, higher frequencies of SCD crises and hospitalization have been reported in other African countries, such as Cameroon^5,6,27,37^. Given The Gambia’s geographic proximity to Senegal and the presence of shared ethnic groups, it is plausible that similarities in β-globin gene cluster haplotypes may contribute to the relatively milder clinical phenotypes observed in the Gambia. The Senegal and Indian β-globin classical haplotypes have been associated with elevated levels of HbF, which is known to inhibit HbS polymerization and mitigate disease severity (Bitoungui et al., 2015; Diallo et al., 2024; Esoh & Wonkam, 2021).

While we observed low prevalence of SCD-related chronic complications, the variation in chronic complications between cohorts is also profound. While leg ulcers, priapism, and stroke predominated in rural settings, osteomyelitis and heart disease were more common in the urban cohort. These may reflect differences in lifestyle, environmental factors, healthcare access, or divergent clinical practices in diagnosis and record-keeping ^5,6,27–29,38^. However, both cohorts operate under similar leadership and use comparable electronic database systems to detailed clinical data of the patients.

One of the most significant contextual findings in the study is no documented evidence of hydroxyurea use in the medical records of both cohorts in The Gambia, consistent with limited use in many African countries^39^. Despite this, supportive therapies such as folic acid, penicillin, and analgesics were widely administered. In our study, we observed that folic acid supplementation was positively associated with haemoglobin levels, while penicillin prophylaxis was linked to reduced infection rates and, indirectly, fewer SCD crises. These findings reinforce the continued importance of basic supportive care in settings where disease-modifying therapies such as hydroxyurea remain constrained. In CSSD cohort, which described the natural history of SCD prior to the implementation of routine prophylaxis of penicillin as part of standard of care in SCD management, bacteraemia was the leading cause of mortality in under five SCD children^28^. In contrast, our study recorded no SCD-related mortality in the first five years of life, coupled with no infection-related mortality in the rural cohort, a finding that could be attributed to the widespread penicillin use, comprehensive malaria prophylaxis use, and routine folic acid supplementation among Gambian children with SCD.

Our multivariable analyses yielded several important findings. Firstly, age of diagnosis and gender were not significant predictors of crises frequency or major clinical outcomes; however, age was found to be indirectly associated with SCD crises and transfusions in the urban cohort with no obvious pattern in the rural cohort. This observation was in contrasts with findings on pain rate in the longitudinal analysis of patients with at least six years of follow up in CSSD cohort, which showed pain rate increase with age for up to 30 years and remain relatively stable thereafter ^40^. It is possible that there is improved disease self-management or awareness among older SCD patients in The Gambia, which will need further exploration via well designed social science approach. Interestingly, seasonality which is frequently linked to SCD crises patterns in other settings ^10,41^ did not strongly predict outcomes, though temporal trends in SCD crises episodes appeared to loosely follow humidity distribution patterns in the urban cohort. This pattern warrants further investigation, ideally using prospective study designs. Further, our multivariate analysis revealed no gender predisposition for the rate of SCD crises and hospital utilisation (hospitalisation and clinic visits) which aligns with findings reported in Senegal and Nigeria^18,26,42^.

Several limitations of this study must be acknowledged. The retrospective data retrieved, while allowing for the inclusion of a broad patient population, had missing or inconsistently documented clinical data in medical records. Such data gaps may have attenuated the observed associations between important non-genetic correlates such as age at diagnosis and seasonality, and SCD clinical outcomes. Moreover, reliance on electronic medical records and use of ICD-10 codes may have led to underestimation of some SCD-related complications incidence, none-reported cases of early SCD-related mortality in both cohorts, and under/over-estimation of clinic visits, sickle cell crises, and hospitalisation. Another important limitation is the absence of important HbF biomarker and comprehensive classical haplotype data, which restricts our ability to fully elucidate the underlying mechanisms contributing to the relatively mild clinical SCD phenotypes observed in The Gambia. The retrospective nature of our data could have contributed to a very wide confidence interval observed the association studies of supportive treatments effects on clinical outcomes. Despite these limitations, our findings provide a valuable baseline for future longitudinal and genomic studies aimed at characterizing the natural history and genetic background of SCD in this population

This study highlights the predominance of relatively mild clinical phenotypes of SCD and a low prevalence of SCD-related complications in The Gambia, alongside delayed diagnosis and the absence of hydroxyurea use. These findings underscore the critical role of essential supportive care—such as folic acid supplementation, penicillin prophylaxis, and infection prevention—in improving the clinical outcomes of individuals with SCD. To strengthen SCD management in The Gambia, it is imperative to implement early diagnostic strategies through universal newborn screening programmes and improve affordable access to hydroxyurea as a disease-modifying therapy. Furthermore, additional studies are warranted to investigate genetic modifiers of disease severity, including variants associated with persistently elevated HbF levels, as well as the influence of sociodemographic, environmental, and clinical factors on disease progression and patient outcomes.

## Supporting information

ethics letter

## Acknowledgements

We gratefully acknowledge the invaluable contributions of the clinical laboratory and Keneba clinical teams, whose efforts enabled access to genotype data essential for confirming the sickle cell disease status of the study population. We also extend our sincere appreciation to the data management team for their critical role in extracting and curating the clinical and epidemiological data from the electronic databases maintained by the Medical Research Council Unit The Gambia at the London School of Hygiene and Tropical Medicine. Importantly, the Science for Africa Foundation Deltas II programme and West Africa Centre for Cell Biology and Infectious pathogens for the funding. We acknowledged professor Nuredin Mohammed for his guidance and review of the study analysis plan, and model selection for the multivariate analysis.

## Author Contributions

M.D. conceived the study, performed the data analysis, and drafted the initial manuscript. M.D., K.E., A.A., C.C., and L.A. contributed to study conceptualisation, interpretation of the findings, and critical revision of the manuscript. K.F., B.S., L.M., and B.S. supported data extraction and contributed to interpretation of the results. E.O. and N.I.C. assisted with data analysis and manuscript preparation. All authors reviewed, revised, and approved the final manuscript.

## Data Availability

The source data underlying the graphs and tables in this manuscript are provided as Supplementary Data 1 (MRCG Fajara Cohort) and Supplementary Data 2 (MRCG Keneba Cohort).

