## Supplementary file for "Sickle Cell Disease Demographics and Clinical Epidemiology in Gambian Urban and Rural Cohorts Retrospective Analysis"

**Methods**

**Study site description**

**MRCG Fajara clinic (also known as Fajara clinical service department)**
The **MRCG Fajara clinic** site provides general medical and paediatric services to both outpatients and inpatients, as well as primary healthcare for MRCG staff. Core services include outpatient consultations; provision of essential drugs and dressings; diagnosis and treatment of non-surgical conditions in adults and children; endoscopy; radiology (X-ray and ultrasound); and ambulance transfer to referral centres when indicated. The site has fully equipped microbiology, biochemistry, and haematology laboratories, with additional tuberculosis, serology, and molecular diagnostic testing provided through affiliated research laboratories.
Although a small number of long-term patients may receive emergency access when clinically indicated, the facility has a limited range of investigations and treatments. Computed tomography and magnetic resonance imaging, dialysis, high-dependency or intensive care facilities, obstetric, gynaecological, surgical, and psychiatric services are not provided on site, and patients requiring such care are referred to external specialists. The unit does not operate a 24-hour emergency service for the general public; patients presenting outside normal working hours are redirected to alternative facilities(MRC Unit The Gambia, 2017).

**MRCG Keneba Clinic (West Kiang)**
The Keneba clinic delivers primary healthcare to the local population of West Kiang. Three physicians supported by nurses and midwives provide mainly outpatient care for general medical and paediatric conditions during morning clinics. A limited midwifery service offers antenatal care and routine deliveries, and a two-bedded ward is available for short admissions. Unlike the Fajara site, Keneba provides a 24-hour emergency service for the public. Patients with conditions beyond the clinic’s scope are referred to Bwiam General Hospital, Edward Francis Small Teaching Hospital, or MRCG Fajara clinical service department. All services at Keneba are free of charge for residents of West Kiang; patients from outside the district are issued prescriptions to purchase medications from private pharmacies(Hennig et al., 2015).

**Results**

**Table S1:** Proportions of SCD patients registered in MRCG Keneba Clinic with reported chronic complications

| **Chronic complications** | **Number of SCD patients (%)** | **Percentage** |
| --- | --- | --- |
| **No Complication reported** | **100(64.94** | **64.94** |
| **Leg Ulcer** | **15** | **9.74** |
| **Malnutrition** | **9** | **5.84** |
| **Stroke** | **5** | **3.25** |
| **Heart disease** | **4** | **2.60** |
| Arthritis | 3 | **1.95** |
| **Hypertension** | **3** | 1**.95** |
| Kidney disease | 2 | 1.30 |
| Splenomegaly | 2 | 1.30 |
| Depression | 1 | 0.65 |
| Epilepsy | 1 | 0.65 |
| Epistaxis | 1 | 0.65 |
| Eye Complication (conjunctivitis) | 1 | 0.65 |
| Stunted growth | 1 | 0.65 |
| Subfertility | 1 | 0.65 |

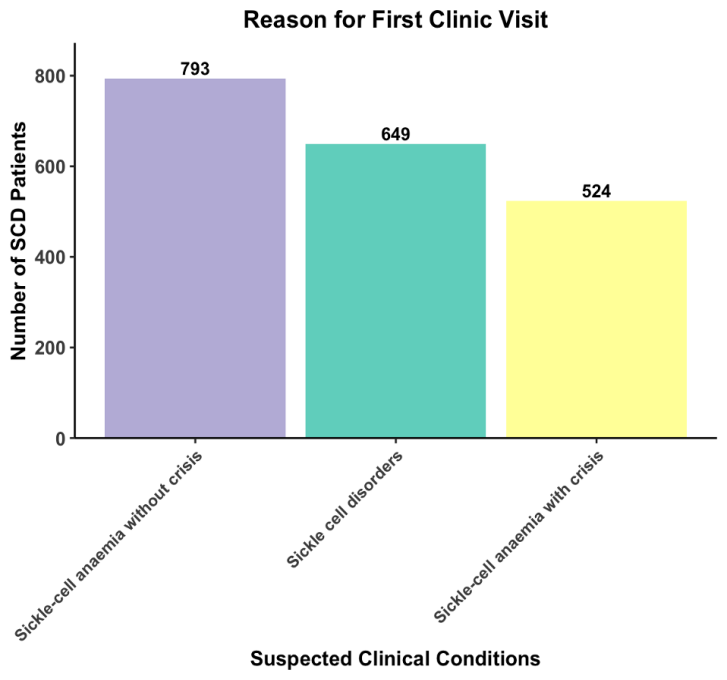

**A**

**B**

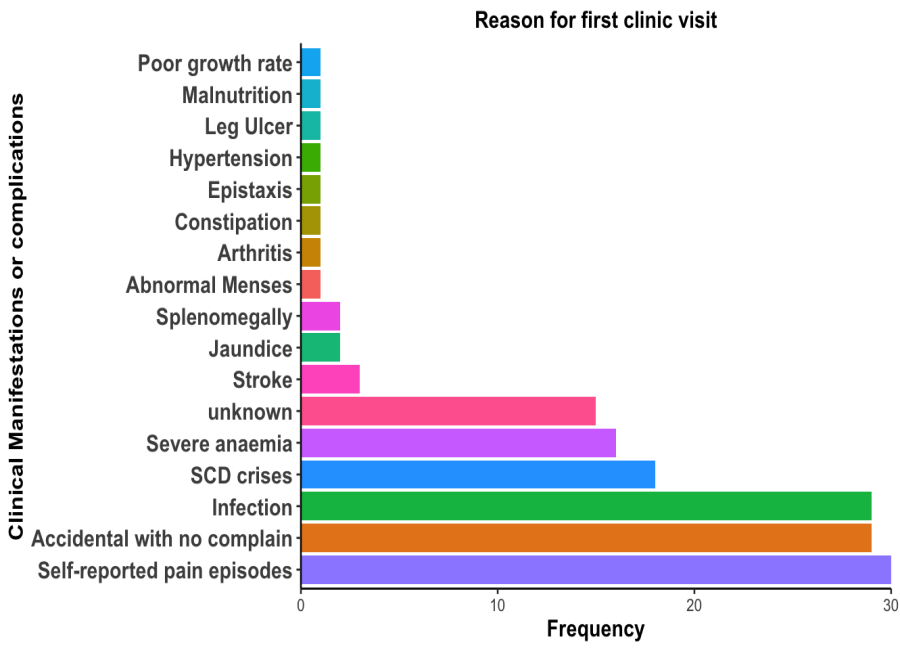

**Figure S1:** Reason for first clinical visit among SCD patients. A) MRCG keneba clinic data B) MRCG Fajara clinic data.

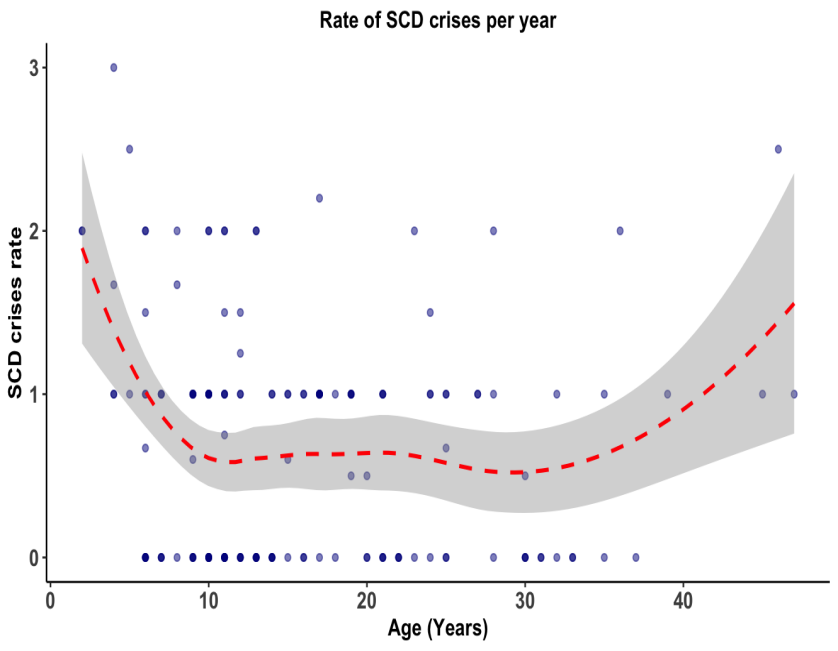

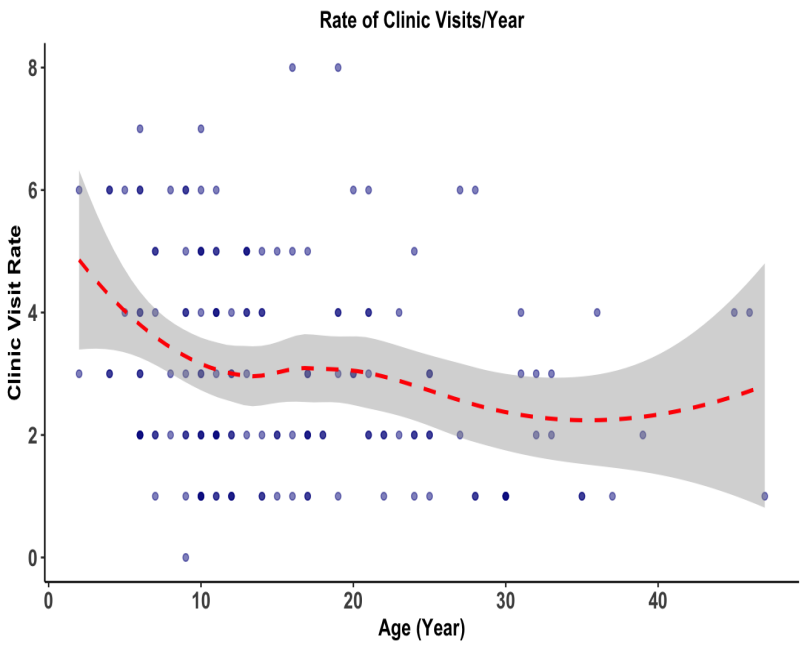

**Figure S2:** Correlation between SCD crises and clinic visit rate, and the age in the two cohorts using the combined dataset.

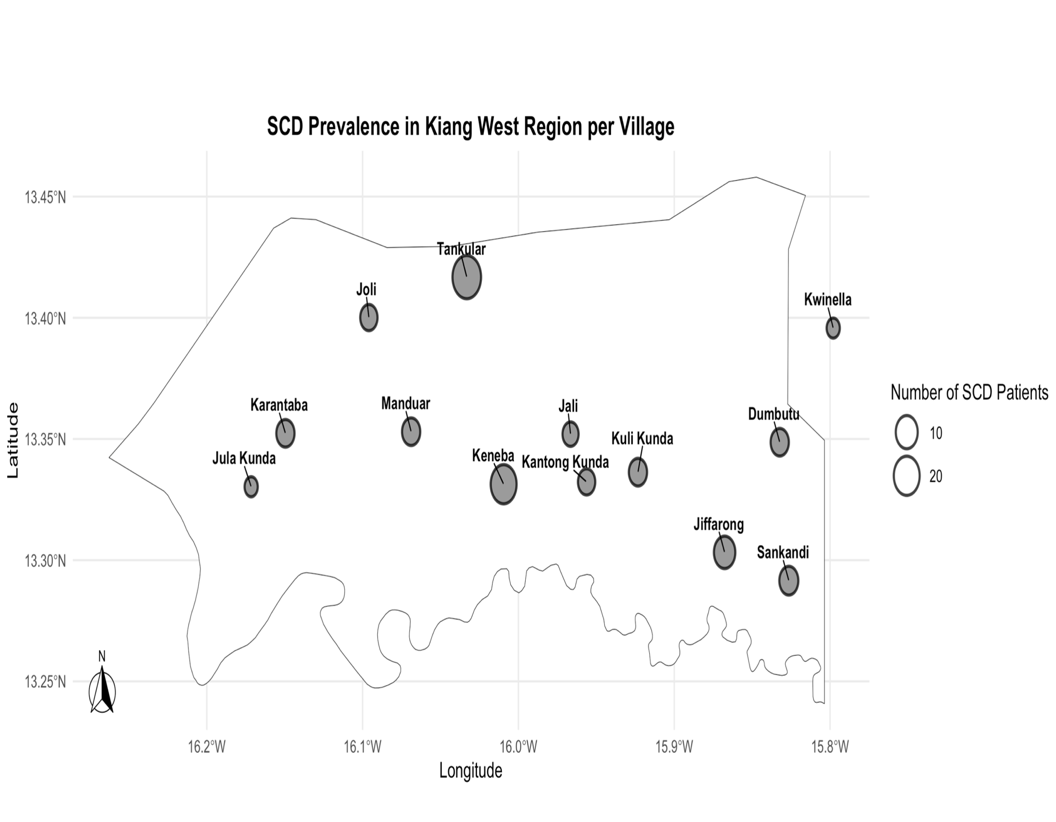

**A**

B

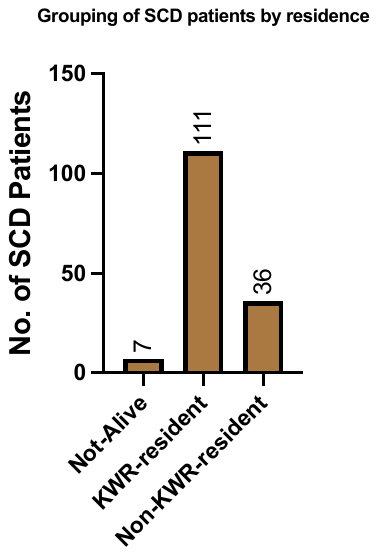

**Figure S3:** Residential status of SCD patients and their proportions across all the villages in the Kiang West Region (KWR) of The Gambia for rural cohort. A) Grouping SCD patients by residence B) Grouping SCD patients by village

**Table S3:** Comprehensive list of acute and chronic SCD-related complications seen in the dataset of SCD patients visiting MRCG Fajara Clinic (urban cohort) with their frequencies and proportions

| **Complications** | **Frequencies (%)** |
| --- | --- |
| Sickle cell anaemia with crises | 122(49.59) |
| SCD crises with infection | 34(13.82) |
| Unknown* | 20(8.13) |
| SCD crises with severe anaemia | 12(4.88) |
| SCD crises with arthritis | 7(2.85) |
| SCD crises with heart disease | 7(2.85) |
| SCD crises with osteomyelitis | 5(2.03) |
| SCD crises with malaria | 4(1.63) |
| Acute lower respiratory infection | 3(1.22) |
| Osteomyelitis | 3(1.22) |
| Pneumonia | 3(1.22) |
| Rheumatic heart disease | 3(1.22) |
| SCD crises with FUO | 2(0.81) |
| SCD with gastrointestinal complication | 2(0.81) |
| Sepsis | 2(0.81) |
| Severe anaemia | 2(0.81) |
| SCD crises with Gastritis | 1(0.41) |
| SCD crises with asthma | 1(0.41) |
| SCD crises with liver disease | 1(0.41) |
| SCD crises with stroke | 1(0.41) |
| Acute chest syndrome | 1(0.41) |
| Acute nephritic syndrome | 1(0.41) |
| Arthritis | 1(0.41) |
| Asthma | 1(0.41) |
| Bacterial pneumonia | 1(0.41) |
| Female pelvic inflammatory disease | 1(0.41) |
| Gastroenteritis and colitis | 1(0.41) |
| Pain (abdominal and pelvic pain) | 1(0.41) |
| Severe anaemia+pneumonia | 1(0.41) |
| Skin infections | 1(0.41) |
| Vitamin b12 deficiency anaemia | 1(0.41) |

**Admitted with unknown complication*

**Table S4:** Summary of clinical outcomes of SCD patients found in both Kiang West region and MRCG Fajara clinic cohorts

| Characteristics | Age  0-5 | Age  5-10 | Age  10-18 | Age  18-30 | Age  30-50 | Age  >50 | Male | Female |
| --- | --- | --- | --- | --- | --- | --- | --- | --- |
| **MRCG Fajara Cohort** | | | | | | | | |
| # clinic visits/pt | 6.44  (889/138) | 8.13  (1682/207) | 9.07  (2315/255) | 7.11  1088/153 | 6.92  (436/63) | 18.25  (73/4) | 7.92  (3372/426) | 7.89  (3111/394) |
| # visits for sickle cell anaemia with crises /pt | 1.05  (117/11) | 1.02  (187/184) | 1.24  (286/230) | 0.96  (131/136) | 1.47  (85/57) | 8.5  (17/2) | 1.11  (419/376) | 1.17  (404/344) |
| # visits which resulted admission in MRCG ward | 1.91  (63/33) | 1.86  (69/37) | 1.84  (68/37) | 1.2  (18/15) | 2.25  (27/12) | 2.0  (2/1) | 1.91  (141/74) | 1.74  (106/61) |
| # whole blood transfusion due to Severe anaemia/pt | 1.33  (32/24) | 1.12  (19/17) | 1.0  (9/9) | 1  (4/4) | 1.00  (3/3) | 0 | 1.16  (43/37) | 1.20  (24/20) |
| Mortality (n=none) | - | - | - | - | - | - | - | - |
| **MRCG KWR Cohort** | | | | | | | | |
| # clinic visits/pt | 5.75  (305/53) | **8.73**  (454/52) | **12.52**  (801/64) | **11.31**  (441/39) | 2.33  (14/6) | 1  (1/1) | 11.44  (904/80) | 15.22 (1111/74) |
| # SCD crises/pt | 1.76  (37/21) | **2.79**  (92/33) | **3.81**  (137/36) | **4.71**  (80/17) | 1.0  (2/2) | - | 4.40  (176/40) | 4.52  (172/38) |
| # visits which resulted in overnight admission in Keneba | 1.33  (4/3) | 1.67  (5/3) | 1  (4/4) | - | - | - | 2  (6/3) | 1.4  (7/5) |
| #referrals for transfusion/pt | 1  (3/3) | 1  (2/2) | 1  (4/4) | 1  (1/1) | 0 | 0 | 1(3/3) | 1(7/7) |
| # visits for infection/pt | 2.21  (62/28) | **2.42**  **(63/26)** | **3.03**  **(105/35**) | **3.36**  **(64/19)** | 1.50  (3/2) | 0 | 3.45  (131/38) | 4.28  (167/39) |
| # visits for severe anaemia/pt | 1  (8/8) | 1  (11/11) | **1.37**  **(26/19)** | **1.25**  **(10/8)** | 0 | 0 | 1.30  (26/20) | 1.38  (29/21) |
| Mortality (n=7) | - | 1(14.3) | 1(14.3) | 4(57.1) | - | 1(14.3) | 5(71.4) | 2(28.6) |

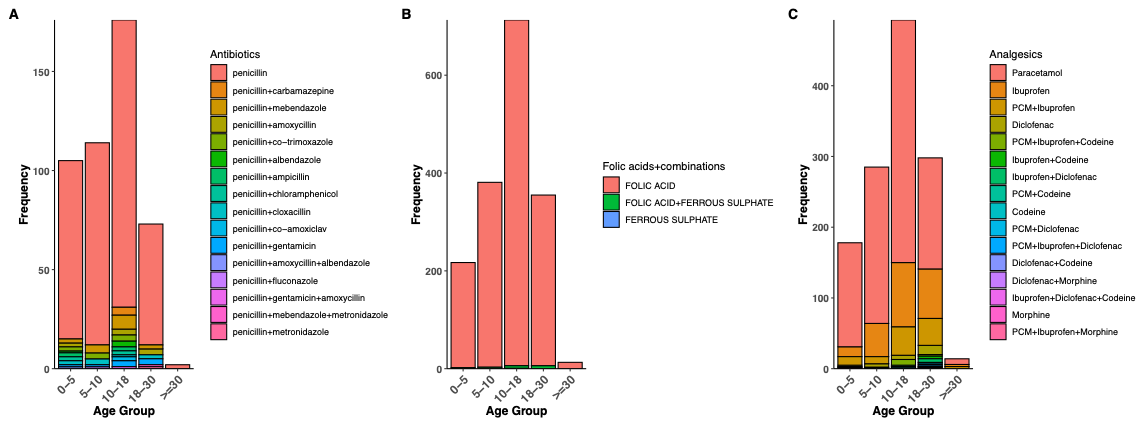

**Figure S4:** **Routine medications prescribed at the MRCG Keneba clinic across different age groups of SCD patients, illustrating the distribution of commonly used treatments**. (A) Use of penicillin and penicillin-based combination antibiotics, showing that penicillin was prescribed across all age groups. (B) Prescriptions for folic acid and its combination with ferrous sulphate.(C) Prescriptions for analgesics used in pain management, indicating the diversity of pain medications administered.

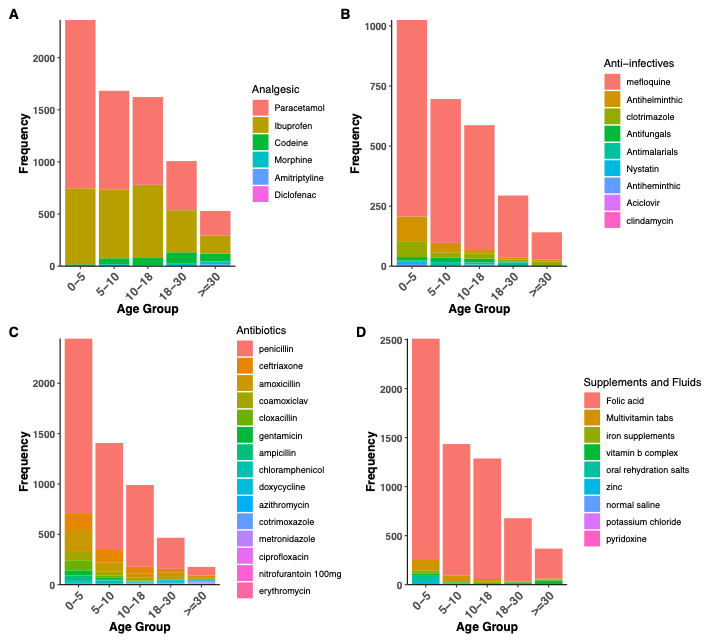

**Figure S5:** **Routine medications prescribed at the MRCG Fajara clinic across different age groups of SCD patients. A)** Prescriptions for analgesics used in pain management, indicating the diversity of pain medications routinely administered to SCD patients**.** B) Anti-infectives, medications prescribed for infection prevention or treatment. C) Wide range of antibiotics used including penicillin. D) Supplements and fluids prescribed for SCD patients

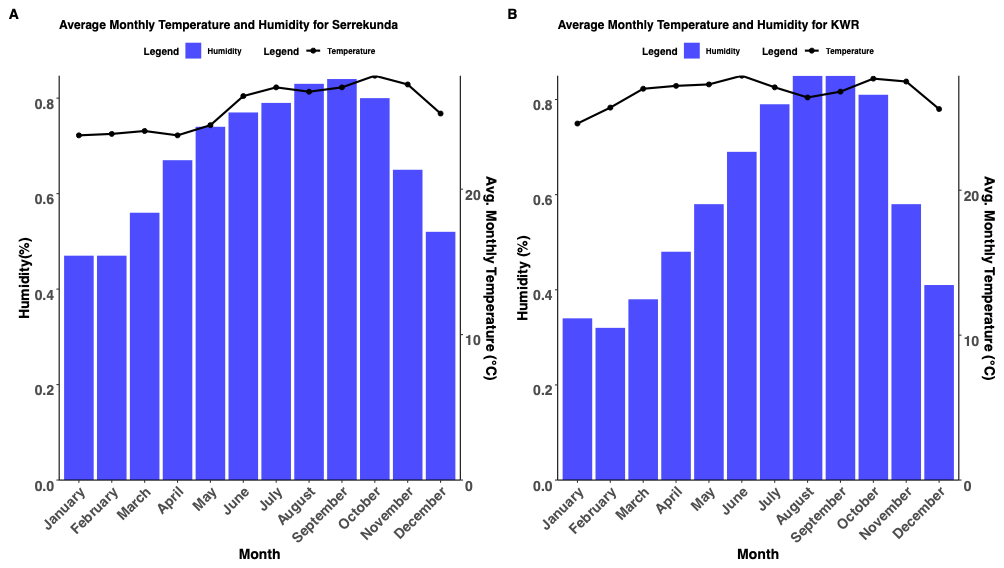

**Figure S6: Monthly distribution pattern of temperature and humidity extracted from world** . **A)** humidity and temperature pattern for lower river region including Kiang West Region **B)** humidity and temperature pattern for Kanifing municipal council, Serekunda. The temperature and humidity data of the Gambia, specifically lower river region and Kanifing municipal council, extracted from World Bank Group climate knowledge portal span 1991 to 2020 (World Bank Group, 2025).

World Bank Group 92025) Climate knowledge portal. Available at: <https://climateknowledgeportal.worldbank.org/country/gambia/climate-data-historical>
