## Supplementary material for "Sickle Cell Disease Demographics and Clinical Epidemiology in Gambian Urban and Rural Cohorts Retrospective Analysis": ethics letter

C/o MRC Unit: The Gambia @ LSHTM, Fajara
P.O. Box 273, Banjul
The Gambia, West Africa


The Gambia Government/MRC Joint

### ETHICS COMMITTEE

Mr. Mustapha Dibbasey
MRCG at LSHTM

7 February 2025

Dear Mr. Mustapha,
**Study Title:** Descriptive Analysis of Existing Medical Records of Sickle Cell Disease Patients Registered at all MRCG at LSHTM Clinics: A Retrospective Study.

**Project ID/ethics ref:** 31114

Thank you for submitting your application which was considered by the Gambia Government/MRCG Joint Ethics Committee at its meeting held on Thursday 30 January 2025.

**Confirmation of ethical opinion**

On behalf of the Committee, I am pleased to confirm a favourable ethical opinion for the above research on the basis described in the application form, and supporting documentation.

**Approved documents**

The final list of documents reviewed and approved by the Committee is as follows:

| Document Type | File Name | Date | Version |
|---|---|---|---|
| Research Ethics training certificate | Research_Ethics_online_training_certificate | 09/12/2024 | 1 |
| Protocol / Proposal | Mustapha_Dibbasey_Objective_1_SCC | 20/12/2024 | 1 |
| Protocol / Proposal | Infographic clinic_FINAL October 2022 | 20/12/2024 | 1 |
| Protocol / Proposal | Infographic_Keneba_Biobank_for patients | 20/12/2024 | 1 |
| Investigator CV | Prof Alfred CV | 09/12/2024 | 1 |
| Investigator CV | Mustapha_Dibbasey_CV_Latest | 09/12/2024 | 1 |
| Covering Letter | Cover_Letter_Response_SCC | 20/01/2025 | 1 |

**After ethical review**

The Principal Investigator (PI) or delegate is responsible for informing the Ethics Committee of any subsequent changes to the application. These must be submitted to the Committee for review using an Amendment Form. Amendments must not be initiated before receipt of written favourable opinion from the Committee.

The PI or delegate is also required to notify the Ethics Committee of any protocol violations and/or Suspected Unexpected Serious Adverse Reactions (SUSARs) which occur during the project by submitting a Serious Adverse Event form. An annual report should be submitted to the Committee using an Annual Report Form on the anniversary of the approval of the study during the lifetime of the study. At the end of the study, the PI or delegate must notify the Committee using an End of Study Form.

All aforementioned forms are available on the ethics online applications website and can only be submitted to the committee via the website at: http://leo.lshtm.ac.uk. Additional information is available at: www.lshtm.ac.uk/ethics.

With best wishes

Yours sincerely

MKCham

**Dr Mohammadou Kabir Cham**
**Chairperson, Gambia Government/MRCG Joint Ethics Committee**

**C/O MRC Unit The Gambia at LSHTM**
**PO Box 273 Banjul, The Gambia**
**West Africa**
**Switchboard (+220) 4495442/6 Ext 2308**
**Fax (+220) 4495919/4496513**
****
Intranet: http://mrcportal/Committees/SCC/SitePages/Home.aspx
Webpage:https://mrcportal.mrc.gm/Committees/SCC/SitePages/Home.aspx
